# Algorithmic Fairness and Bias Mitigation for Clinical Machine Learning: A New Utility for Deep Reinforcement Learning

**DOI:** 10.1101/2022.06.24.22276853

**Authors:** Jenny Yang, Andrew A. S. Soltan, David A. Clifton

**Affiliations:** Institute of Biomedical Engineering, Dept. Engineering Science, University of Oxford; John Radcliffe Hospital, Oxford University Hospitals NHS Foundation Trust; RDM Division of Cardiovascular Medicine, University of Oxford

## Abstract

As machine learning-based models continue to be developed for healthcare applications, greater effort is needed in ensuring that these technologies do not reflect or exacerbate any unwanted or discriminatory biases that may be present in the data. In this study, we introduce a reinforcement learning framework capable of mitigating biases that may have been acquired during data collection. In particular, we evaluated our model for the task of rapidly predicting COVID-19 for patients presenting to hospital emergency departments, and aimed to mitigate any site-specific (hospital) and ethnicity-based biases present in the data. Using a specialized reward function and training procedure, we show that our method achieves clinically-effective screening performances, while significantly improving outcome fairness compared to current benchmarks and state-of-the-art machine learning methods. We performed external validation across three independent hospitals, and additionally tested our method on a patient ICU discharge status task, demonstrating model generalizability.

## 1 Introduction

Ever-increasing computational resources, paired with the availability of exponentially-rising amounts of digital health data, is transforming our understanding of both general human health and personalized health assessment. Although the benefits of using machine learning (ML)-based technologies are clear, a large obstacle to implementation is gaining clinicians’ and patients’ trust in its efficacy and reliability. Thus, when developing data-driven tools, careful consideration needs to be given to the fairness and equity of models, especially in healthcare contexts, as algorithmic findings can directly impact clinical decision-making and patient care.

It is well-known that machine learning models are susceptible to biases present in training data. If a model unintentionally learns these biases during training, it may be unable to capture the true relationship between a set of features and the related target outcome. This can lead to unfair decision-making, which we define as any outcome that is skewed towards a particular group or population [1]. Although this issue is relevant in many disciplines, we focus on clinical applications for three reasons - 1) a biased model can lead to poorer performance and inaccurate predictions for critical and, potentially, life-altering decisions; 2) a bias against particular groups can result in those patients receiving poorer care compared to those in other groups; and 3) a biased model can exacerbate and propagate existing inequities in healthcare/society. All of these can compromise clinician and patient trust, making it more difficult to deploy machine learning models into clinical practice.

One type of inequity that has frequently been discussed within healthcare, is ethnicity. And specifically, in the context of data-driven algorithms, ML models have been shown to be susceptible to ethnicity-based biases. For example, a previous study found that a model used to predict recidivism was biased against black defendants, falsely labeling them as future criminals at almost twice the rate as white defendants [2]. With respect to clinical applications, researchers have similarly found that machine learning models perform unequally across different patient populations [3], and that outcomes can negatively impact those in underrepresented groups [4]. This is especially relevant in healthcare, as different regions may be heavily skewed towards certain ethnic groups. For example, randomized trials evaluate treatment effects for a trial population; however, participants in clinical trials are often demographically unrepresentative of the patient population that ultimately receives the treatment [3,5]. Thus, if a model was designed to determine who to give a certain drug or intervention to, minority groups (such as ethnic minorities, women, obese patients, etc.) might receive the least, further deepening demographic inequities within healthcare. In addition to poorer health and treatment outcomes, this issue is also relevant with respect to privacy preservation and statistical disclosure, as some regions may have a very small number of patients of a given ethnicity; and thus, if a machine learning model is biased against this group, there is an increased probability of identifying these patients [6].

Another area where inequity can exist is between different healthcare centres. Previous studies have found that health outcomes and healthcare practice can vary widely across regions and between hospitals. These include variations in disease prevalence/mortality, quality of healthcare services, and specific devices used (e.g. blood analysis devices) [7,8,9,10,11]. Thus, a model created using real-world data from one hospital may not generalised to a new setting, as the methods used to collect, process, and organize data may have unintentionally encoded site-specific biases [12]. To address this, many ML-based clinical projects have tried to combine datasets from multiple hospitals to increase the amount of training data available, as achieving generalizability typically requires large amounts of data. However, since different centres may have varying amounts of training data available, models can still accumulate site-specific biases during training. If these biases become reflected in a model’s decisions, then certain hospitals may be isolated for exhibiting poorer outcomes [6], which can both widen inter-hospital inequities and discourage some hospitals from implementing ML-based technologies.

The current literature for addressing bias mitigation at the algorithmic-level has primarily been focused on adversarial debiasing - a technique where a model is trained to learn parameters that do not infer sensitive features. Here, a predictor network is trained against an adversary network, where the adversary assures that the predictor’s output is not correlated with the specified sensitive feature (i.e., the unwanted bias which we are trying to mitigate). This technique has been used to develop models that output fair predictions, and has previously been successful in reducing gender (male versus female) bias in salary prediction [13,14] and ethnicity (black vs white) bias in recidivism prediction [15]. Adversarial models have also been used to effectively predict COVID-19, whilst simultaneously improving outcome fairness with respect to site-specific (hospital) and ethnic biases [6]. As specialized methods have been shown to be necessary for mitigating unwanted biases, we aimed to use a reinforcement learning framework (instead of an adversarial one) to optimize fairness outcomes.

Reinforcement learning (RL) - whereby an agent interacts with an environment to learn a task - has been linked to many real-world artificial intelligence (AI) successes, with many well-known exemplars in game play and control. However, the core elements of RL have been shown to be successful on a wider range of tasks, including those which, on the surface, do not appear to have a particular “agent” interacting with an “environment” (which is typically regarded as the standard RL set-up [16,17]). Such problems include classification tasks, which have commonly been addressed using standard supervised learning algorithms (where an input is mapped through a model to predict a class label). RL instead, uses an agent to interact with the input to determine which class it belongs to, and then receives an immediate reward from its environment based on that prediction. A positive reward is given to the agent when a label is correctly predicted, and a negative one is given otherwise. This feedback helps the agent learn the optimal “behavior” for classifying samples correctly, such that it accumulates the maximum rewards. To do this, an agent performs actions that set memory cells, which can then be used by the agent (together with the original input) to select actions and classify samples [18]. Specialized reward functions have previously been successfully used for mitigating large data imbalances with respect to the predicted label [19,20]. Thus, instead of focusing on label imbalance, we aimed to formulate a deep RL framework with the specific purpose of improving algorithmic fairness and mitigating unwanted biases.

To summarize, we introduce a novel reinforcement learning-based framework specifically tailored for mitigating unwanted biases during training. We specifically used a dueling double deep Q-network (DDQN) architecture and evaluated our method on a real-world, clinical task - COVID-19 screening using anonymized electronic health record (EHR) data from hospital emergency rooms. Using this task, we aimed to mitigate any unwanted ethnicity-based and site-specific (hospital) biases. To demonstrate the utility of our method across diverse clinical tasks, we performed additional analyses on a patient discharge status task using EHR data from intensive care units (ICUs). Although we use clinical case studies, the framework introduced can be generalized across many different domains, and can be applied to a variety of tasks and features.

## 2 Methods

We trained models to predict the COVID-19 status of patients presenting to hospital emergency departments (EDs) using routinely collected clinical data typically available within 1 hour of arrival (routine blood tests and vital signs). Previous works have shown that machine learning-based methods can identify patients presenting with COVID-19 up to 90% sooner than Polymerase Chain Assay (PCR) testing, achieving high sensitivities and performing effectively as a rapid test-of-exclusion [6,12,21,22]. Additionally, one study has shown that adversarial models were effective at screening for COVID-19 whilst being able to mitigate biases for selected sensitive features [6]. We aimed to build on these existing works, formulating a deep reinforcement learning framework (with a specialized reward function) to effectively screen for COVID-19, while simultaneously mitigating unwanted biases.

### 2.1 Datasets and Pre-processing

To train and validate our models, we used clinical data with linked, deidentified demographic information for patients presenting to EDs across four independent United Kingdom (UK) National Health Service (NHS) Trusts – Oxford University Hospitals NHS Foundation Trust (OUH), Portsmouth Hospitals University NHS Trust (PUH), University Hospitals Birmingham NHS Trust (UHB), Bedfordshire Hospitals NHS Foundations Trust (BH).

With scalability and speed in mind, we trained models using laboratory blood tests and vital signs, as these are widely and routinely collected during the first hour of patients attending emergency care pathways in hospitals in middle- to high-income countries [21]. The specific features included are the same as those used in [12] (also similar to those used in [22] and [6]), allowing for comparison. Table 1 summarizes the final features included.

**Table 1:**
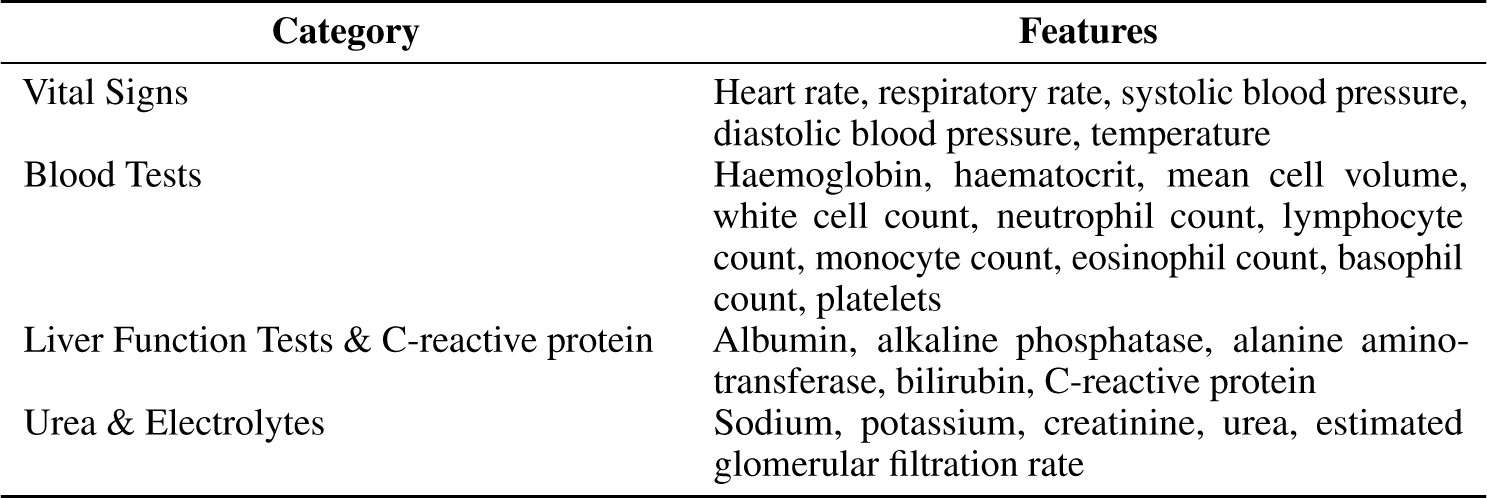
Clinical predictors considered.

| Category | Features |
| --- | --- |
| Vital Signs | Heart rate, respiratory rate, systolic blood pressure, diastolic blood pressure, temperature |
| Blood Tests | Haemoglobin, haematocrit, mean cell volume, white cell count, neutrophil count, lymphocyte count, monocyte count, eosinophil count, basophil count, platelets |
| Liver Function Tests & C-reactive protein | Albumin, alkaline phosphatase, alanine amino-transferase, bilirubin, C-reactive protein |
| Urea & Electrolytes | Sodium, potassium, creatinine, urea, estimated glomerular filtration rate |

For each of the models, a training set was used for model development, hyperparameter selection, and model training; a validation set was used for continuous validation and threshold adjustment; and after successful development and training, three held-out, external test sets were then used to evaluate the performance of the final models. How the data were split are detailed in following sections.

For the training and validation sets used in the ethnicity debiasing model, we used patient presentations exclusively from OUH. From OUH, we curated two data extracts - one from the first wave of the COVID-19 epidemic in the UK (December 1, 2019 to June 30, 2020), and one from the second wave (October 1, 2020 – March 6, 2021) (Supplementary Figure 1). Due to incomplete penetrance of testing during the first wave, and imperfect sensitivity of the polymerase chain reaction (PCR) test, there is uncertainty in the viral status of patients presenting who were untested or tested negative. Thus, from the “wave one” dataset, we only included the positive cases (as determined through PCR tests) in training; and from the “wave two” dataset, we included both positive COVID-19 cases (by PCR) and negative controls. This was done to ensure that the label of COVID-19 status was correct during training. Furthermore, to reasonably evaluate classification performance with respect to ethnicity, we removed any presentations where the label for ethnicity was ambiguous, including those labeled as “unknown”, “mixed”, or “other”. This resulted in 18,687 patients used in training and validation, including 2,083 of which were COVID-19 positive. A ratio of 80:20 was used to split the OUH cohort into training and validation sets, respectively. We then performed external validation on three independent patient cohorts from PUH, UHB, and BH (totalling 38,964 admitted patients, including 1,963 of which were COVID-19 positive). From Table 2, we can see that ethnicity is heavily skewed in our training dataset, making it a possible source of bias during training.

**Table 2:** Summary of number of patients, COVID-19 positive cases, and ethnicity distribution for training, validation, and external test set cohorts included in the ethnicity debiasing task.

|  | Training | Validation | External Test |  |  |
| --- | --- | --- | --- | --- | --- |
|  | OUH | OUH | PUH | UHB | BH |
| **n, patients** | 14,949 | 3,738 | 29,103 | 8,730 | 1,131 |
| **n, COVID-19 positive (PCR)** | 1,672 | 411 | 1,478 | 347 | 138 |
| **Ethnicity:** |  |  |  |  |  |
| White (%) | 14,293 (95.6) | 3,574 (95.6) | 28,704 (98.6) | 6,848 (78.4) | 1,024 (90.5) |
| South Asian (%) | 370 (2.5) | 93 (2.5) | 170 (0.6) | 1357 (15.5) | 71 (6.3) |
| Black (%) | 243 (1.6) | 61 (1.6) | 187 (0.6) | 484 (5.5) | 36 (3.2) |
| Chinese (%) | 43 (0.3) | 10 (0.3) | 42 (0.1) | 41 (0.5) | 0 (0.0) |

In addition to debiasing ethnicity, we also demonstrated the utility of our proposed method for debiasing with respect to the hospital that a patient attended. In order to evaluate bias related to hospital location, presentations from multiple sites needed to be present in the training data. Thus, we combined presentations from all hospital cohorts previously described; however, we additionally included the patient presentations with ambiguous ethnicity labels, as we are no longer focusing on mitigating ethnicity-based biases. Using a 60:20:20 split, we separated the data into training, validation, and test sets, respectively, resulting in 58,339 presentations used in training and validation (including 4,245 of which were COVID-19 positive), and 14,585 presentations in the held-out test set (including 1,056 of which were COVID-19 positive). A summary of training, validation, and test cohorts can be found in Table 3.

**Table 3:** Summary of number of patients, COVID-19 positive cases, and hospital case distribution for training, validation, and held-out test set cohorts used in hospital debiasing task.

|  | Training | Validation | Test |
| --- | --- | --- | --- |
| **n, patients** | 43,754 | 14,585 | 14,585 |
| **n, COVID-19 positive (PCR)** | 3,171 | 1,074 | 1,056 |
| **Hospital:** |  |  |  |
| OUH (%) | 14,104 (32.2) | 4,694 (32.2) | 4,760 (32.6) |
| PUH (%) | 22,750 (52.0) | 7,596 (52.1) | 7,550 (51.8) |
| UHB (%) | 6,186 (14.1) | 2,062 (14.1) | 2,045 (14.0) |
| BH (%) | 714 (1.6) | 233 (1.6) | 230 (1.6) |

Consistent with previous studies, we addressed the presence of missing values by using population median imputation, then standardized all features in our data to have a mean of 0 and a standard deviation of 1.

A summary of training, validation, and test cohorts used in each task can be found in Tables 2 and 3. The full inclusion and exclusion criteria for patient cohorts and summary population statistics can be found in Section C of the Supplementary Material.

To further test our framework, we also trained models to predict the discharge status of patients staying in the ICU, using data from the eICU Collaborative Research Database (eICU-CRD) [23]. Here, we also focus on debiasing ethnicity (which again, is heavily imbalanced in the dataset), demonstrating the generalizability of our method on a new, independent clinical task. Details on the dataset, features, and pre-processing steps used for this task can be found in Section D of the Supplementary Material.

### 2.2 Reinforcement Learning for Classification

To formulate classification as a reinforcement learning task, we model our problem in a sequential decision-making format using a finite Markov Decision Process (MDP). We define the MDP using a tuple of five variables (*s, a, p, r*, γ), where: *s* is the state space of the process, *a* is the action that an agent takes, *p* is the transition probability that results from an action, *r* is the reward expected for a given action, and γ is the discount factor used for future rewards.

For a given *N* x *D* dataset, *N* is the total number of samples and *D* is the number of features in each sample. During training, a batch of data is randomly shuffled and presented to the model in order. Here, *p* is deterministic, as the agent moves from one state to the next according to the order of samples in the training data. The features of each sample presented makes up the state, *s*.

The action, *a*, is the prediction the agent makes when presented with a state, *s*. Given a total number of classification labels, *K*, each *a* is selected from one of *K* classes. With respect to COVID-19 classification, *a* = 0, 1, where 0 corresponds to COVID-19 negative cases and 1 corresponds to COVID-19 positive cases.

Because the selection of an action, *a*, does not determine the following sample, *s*, presented to the agent, an alternative dependency must be introduced between *s* and *a*. To achieve this, a training episode is terminated when an agent incorrectly classifies the minority class, preventing any further reward, *r*. This allows for a relationship between *s* and *a* to be learned, especially when there are severe data imbalances between majority (COVID-19 negative) and minority (COVID-19 positive) classes [20].

### 2.3 Defining Reward for Bias Mitigation

The reward, *r*, is the signal evaluating the success of the agent’s selected action. We introduce a specialized function for reward, uniquely formulated for the purpose of mitigating biases of the chosen sensitive feature, *z*. To do this, we separate the reward function into two components – one to help train a strong classifier, and one to debias with respect to the sensitive attribute. Additionally, as the majority of previous studies have exclusively evaluated bias mitigation for binary attributes, we formulated the reward function to be able to debias multi-class attributes. This is especially important in clinical tasks, as a higher degree of granularity is often required since binning values to fewer (i.e. binary) classes may not be biologically relevant (especially when classes are categorical) and is heavily biased on the sample population [6,20]. Thus, to accommodate for class-imbalance for multi-class sensitive features, we make the reward proportional to the relative presence of a class in the data.

A positive reward is given when the agent correctly classifies the sample (as either COVID-19 positive or negative), and a negative reward is given otherwise. If a negative reward is given (i.e. a sample was misclassified), the absolute reward value given is proportional to the relative presence of the label of the sample in the training data. Thus, the absolute reward value of a sample from the minority class is higher than that in the majority class, making the model more sensitive to the minority class. This helps accommodate for label imbalance during training; and since the primary purpose of the model is to effectively classify COVID-19 status, this sensitivity-differential will help the agent learn the optimal behavior for COVID-19 prediction. To consider the sensitive class, *z* (which we aim to debias), we make the absolute reward for the positive case (i.e. when a sample is correctly classified) proportional to the relative presence of each respective *z* label in the training data, accommodating for any class-imbalances present in a multi-class sensitive attribute. Here, the absolute reward value of a sample from a minority *z* class is therefore higher than that from the majority class, making the model more sensitive to minority *z* labels. By performing debiasing on the positively rewarded states, we already know that the sample was correctly classified for the main task, as debiasing would be inconsequential if the model was not clinically effective for use to begin with. The formulation introduced allows for evaluation of both binary and multi-class tasks and sensitive features. Assuming *M* classes in *z*, and using *l*_*k*_ to represent the COVID-19 label, the reward function is formulated as follows:

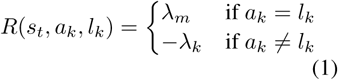

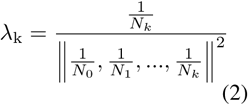

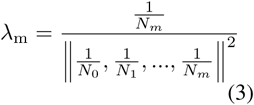

*N* represents the number of instances in class *k* or *m*, and *λ* is a trade-off parameter used for adjusting the influence of each class. We found that models achieved desirable performance when *λ* is the vector-normalized reciprocal of the number of class instances, as shown in Equations 2 and 3. To balance immediate and future rewards, a discount factor, *γ* = 0.1, is used.

#### 2.3.1 Policy Iteration by Q-Learning

An optimal policy *π*\* is a policy that maximizes the expected cumulative reward, *v*^*π*^. This can be interpreted as the value of a state-action combination, and is discovered by iterating through a series of policies, 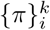, where *π*\*= *π*^*k*^. Using the Bellman equation, *v*^*π*^ can be determined by solving a system of linear equations and calculating a set of *Q*-values, where *Q* represents the action-value function:

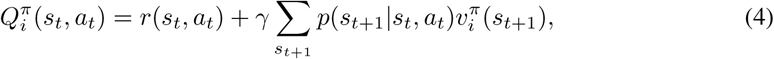

This gives successive policies:

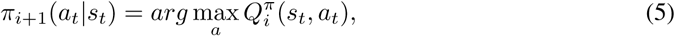

where, *π*^*∗*^ = *arg* max_*a*_ *Q*^*∗*^. Finally, we can use the advantage function, *A*^*π*^, to relate the state-action value function and *Q* function:

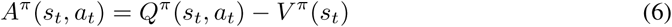

The value function, *v*^*π*^ can be viewed as a proxy for the “goodness” of a particular state, and the *Q*^*π*^ function evaluates the value of selecting a particular action in this state [24]. Thus, *A*^*π*^ can be interpreted as the relative importance of each action.

#### 2.3.2 Dueling Q-Network Architecture

In a typical deep Q-network (DQN) setup, the output layer of the network corresponds to predicted Q-values for state-action pairs. Since only one state-action pair can be trained at a time, it can be difficult to provide update information about the state. To address this, we implement a dueling Q-network, which is capable of training state representations and action representations independent of one another.

For a DQN, the Q-network is implemented as a standard, single-stream neural network, where fully-connected layers are connected in a continuous sequence. The dueling Q-network (Fig. 1), instead, implements a fully-connected neural network with two streams - one for estimating the value (which is scalar) and another to estimate the advantages of each action (which is a vector). These two streams are combined to produce a single output, which is the *Q* function [24].

**Figure 1:**
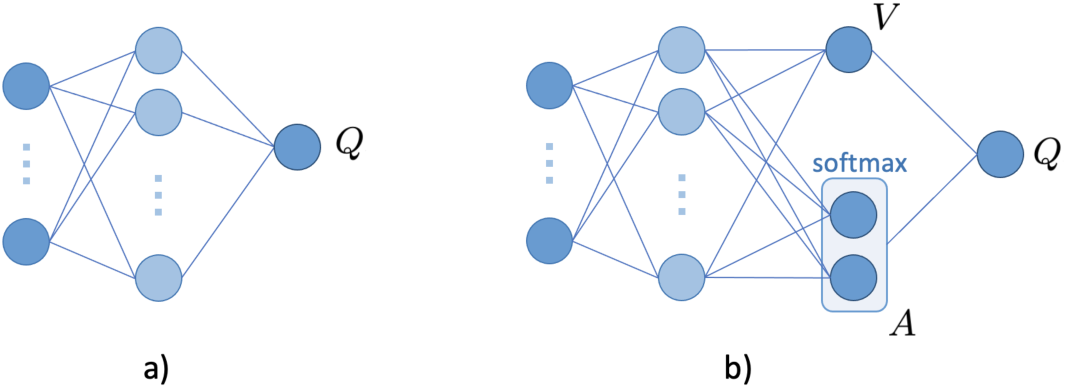
A typical single-stream Q-network is shown in a). A dueling architecture, with two streams to independently estimate the state-values (scalar) and advantages (vector) for each each action is shown in b) (this implements equation 7).

**Figure 2:**
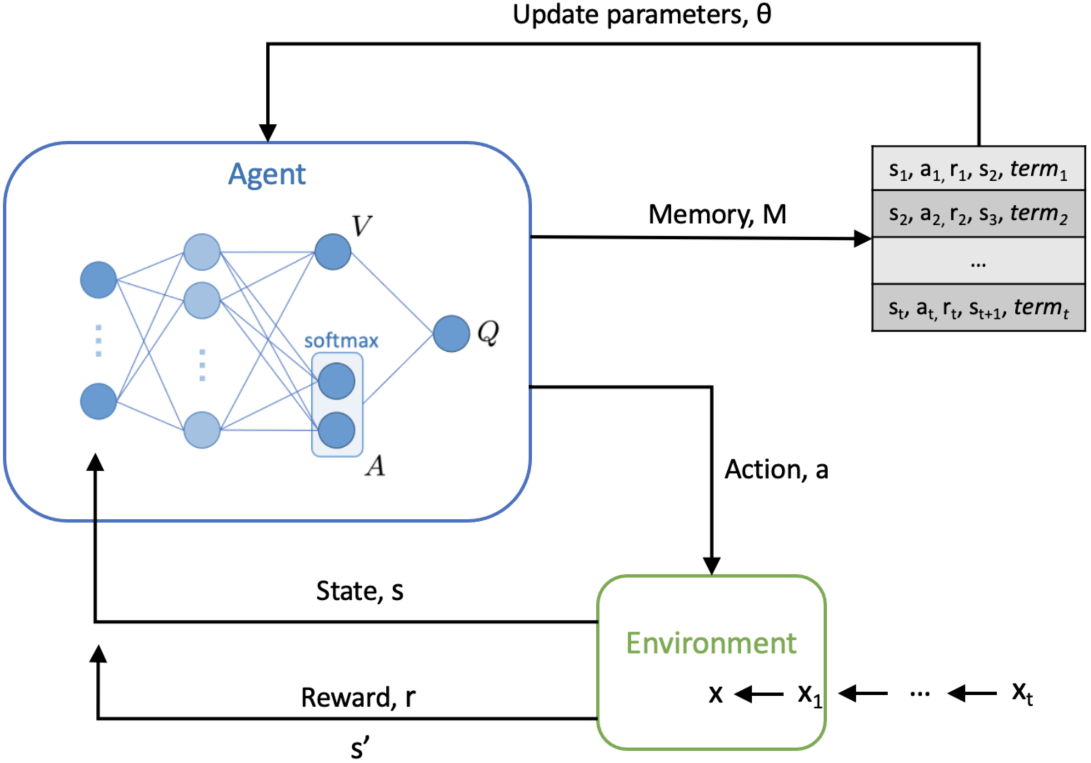
Overview of the reinforcement learning framework used. A dueling network architecture, with two streams to independently estimate the state-values (scalar) and advantages (vector) for each action, is shown.

Based on the definition of the advantage function, we represent *Q* as:

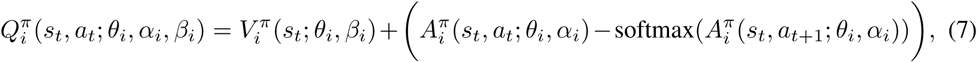

where *α* and *β* represent the parameters of the *A* and *V* streams of the fully connected layers, respectively. The additional softmax module is to allow *Q* to recover *V* and *A* uniquely [24]. Additionally, this extra term does not change the relative rank of *A* (and subsequently, Q-values), which preserves the *ϵ*-greedy policy (which we use in our training).

For the Q-network, we used a fully-connected neural network with one hidden layer, alongside the rectified linear unit (ReLU) activation function and dropout. For updating model weights, the Adaptive Moment Estimation (Adam) optimizer was used during training. We set the exploration probability, *ϵ*, to be linearly attenuated from 1 to 0.01 over the entire training process. Each training period consists of 120,000 steps (i.e. iterations of updating parameters *θ*).

#### 2.3.3 Double Deep Q-Learning

During each episode, combinations of states, actions, and rewards at each step, (*s*_*t*_, *a*_*t*_, *r*_*t*_, *s*_*t*+1_), are saved in the agent’s working memory, *M*. To learn the parameters of the Q-network, *θ*, a randomly sampled subset of these transitions, *B*, are used in the gradient descent step. The mean-squared error loss function is used to optimize the network:

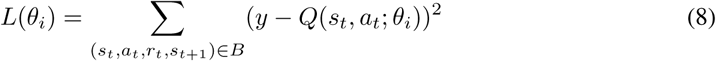

As in standard supervised learning, *y* can be treated as the target to be predicted and *Q*(*s, a*; *θ*_*i*_) as the prediction. We define *y* using the format of a double deep Q-Network (DDQN).

As a standard DQN uses the current Q-network to determine an action, as well as estimate its value, it has been shown to give overoptimistic value estimates [25]. This increases the likelihood of selecting overestimated values (which can occur even when action values are incorrect), making it harder to learn the optimal policy. Thus, double deep Q-Learning (DDQN) was introduced as a method of reducing this overestimation [26]. Unlike a DQN, a DDQN uses the current Q-network to select actions, and the target Q-network to estimate its value [27]. Through decoupling the selection and evaluation steps, a separate set of weights, *θ* ^*′*^, can be used to provide an unbiased estimate of value.

The DDQN algorithm is implemented using the following target function:

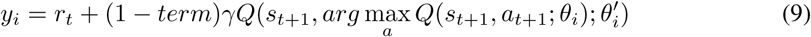

As previously mentioned, a dependency between a state and action needs to be established for the agent to learn a relationship. Thus, within this function, the value of *term* is set to 1 once the agent reaches its terminal state, and 0 otherwise. A terminal state is reached after the agent has iterated through all samples in the training data (or a set number of samples, specified at the beginning of training), or when the agent misclassifies a sample from the minority class (preventing any further reward).

## 3 Reinforcement Learning Training Procedure

In each episode, the agent employs an *ϵ*-greedy behavior policy to select an action, which randomly selects an action with probability *ϵ*, or an action following the optimal Q-function, *arg* max_*a*_ *Q*^*∗*^(*s*_*t*_, *a*_*t*_) with probability 1 − *ϵ*. A subsequent reward is then given from the environment through the process described in Algorithm 1. The overall Q-network is trained according to the DDQN process described in Algorithm 2. The final, optimized Q-network is considered to be the trained classifier.

### Algorithm 1: Environment Reward Procedure

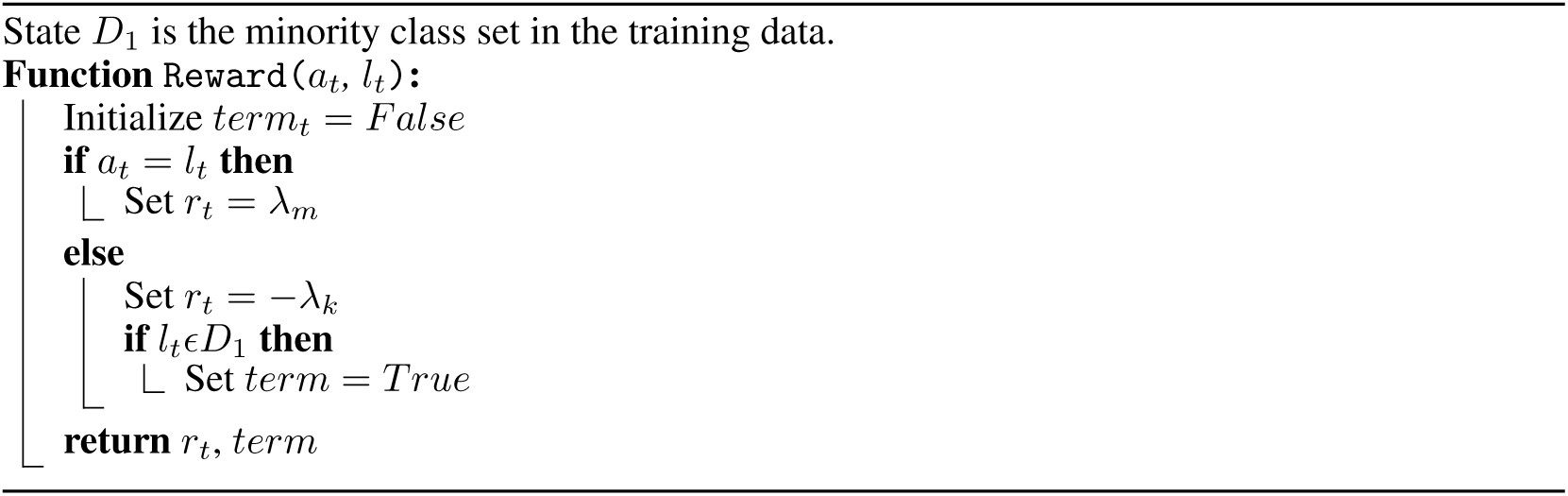

### Algorithm 2: DDQN Training Procedure

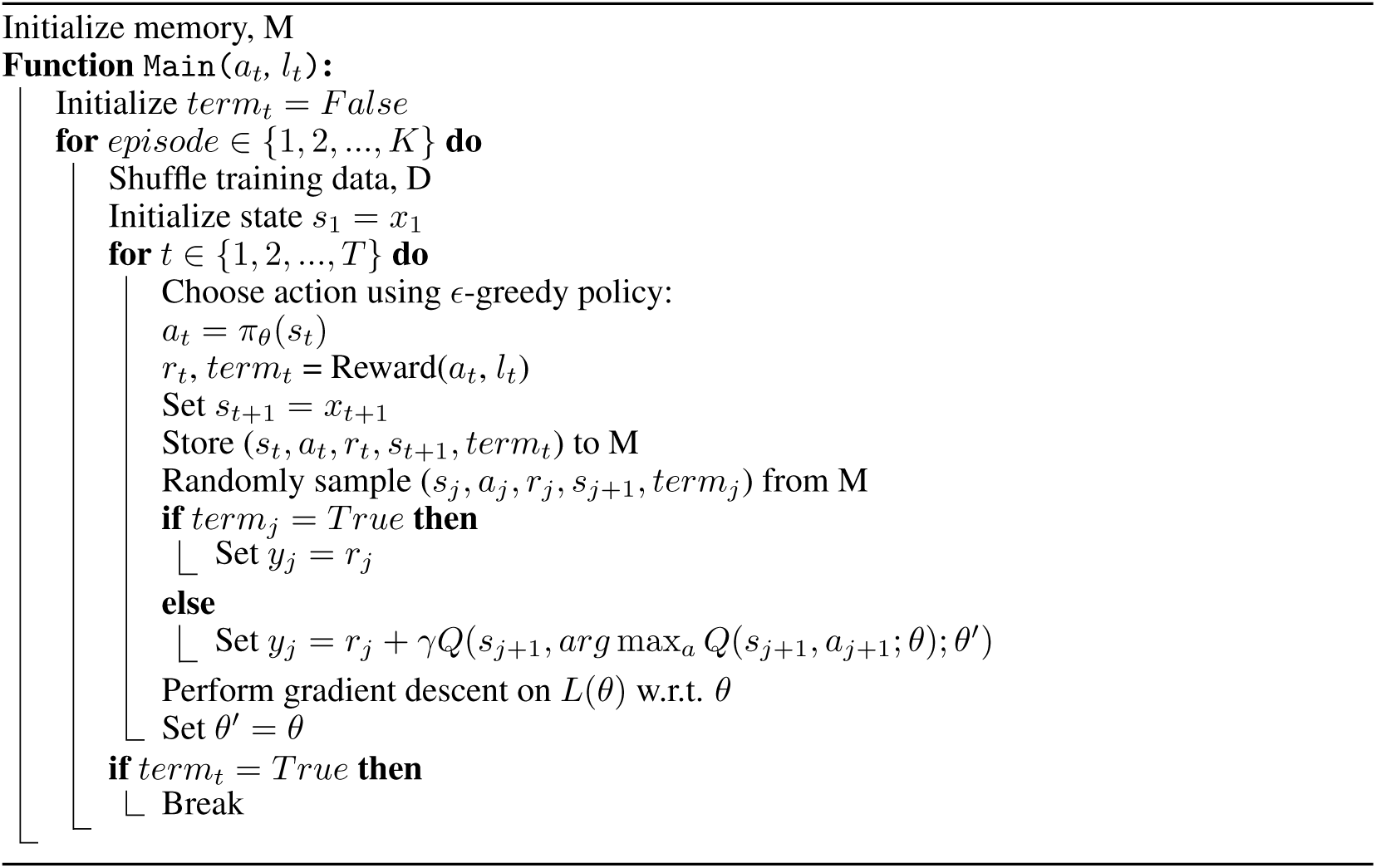

## 4 Model Comparators and Evaluation Metrics

We compare the effectiveness of our method against two baseline models - a fully-connected neural network and XGBoost. XGBoost is a popular ensemble model that has achieved state-of-the-art results on many machine learning challenges, and a standard, fully-connected neural network forms the basis of both an adversarial debiasing framework and our RL-based framework. Thus, comparing our model to these two baselines will ensure that we have trained a strong classifier to begin with. We additionally compare our method to an adversarial debiasing framework, which is the current state-of-the-art method for mitigating biases at an algorithmic level.

Additionally, all comparator methods have previously been shown to be able to effectively screen for COVID-19, using the same datasets, allowing for direct comparison of our method [6,21,22]. Details on network architectures of comparator models can be found in Section B of the Supplementary Material.

To evaluate the performance of COVID-19 prediction, we report sensitivity, specificity, positive and negative predictive values (PPV and NPV), and the area under receiver operator characteristic curve (AUROC), alongside 95% confidence intervals (CIs) based on standard error. CIs for AUROC are calculated using Hanley and McNeil’s method.

As the purpose of our framework is to train models that are unbiased towards sensitive features, we evaluate model fairness using the statistical definition of equalized odds. Here, a classifier is considered fair if true positive rates are equal and false positive rates are equal, across all possible classes of the sensitive attribute [28]. To assess multiple labels (i.e., *>*2), we used the standard deviation (SD) of true positive and false positive scores. SD scores closer to zero suggest greater outcome fairness. The equations used to calculate true positive and false positive SD scores are as follows:

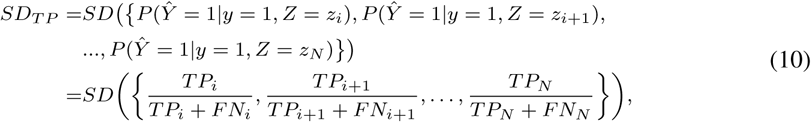

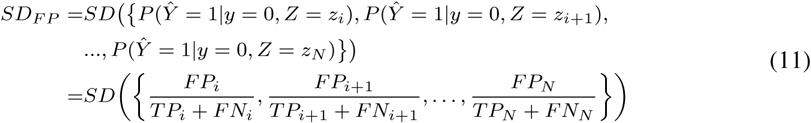

### 4.1 Hyperparameter Optimization and Threshold Adjustment

In each task, appropriate hyperparameter values were determined through grid search and standard 5-fold cross-validation (CV), using respective training sets. Grid search was used to determine: (i) the number of nodes to be used in each layer of the neural network and RL models, (ii) the learning rate, and (iii) the dropout rate. 5-fold CV was used to ensure that hyperparameter values were evaluated on as much data as possible, as to provide the best estimate of potential model performance on new, unseen data. Details on the hyperparameter values used in the final models can be found in Section E of the Supplementary Material.

The raw output of many ML classification algorithms is a probability of class membership, which is then mapped to a particular class. For binary classification, the default threshold is typically 0.5, where values equal to or greater than 0.5 are mapped to one class and all other values are mapped to the other. However, this default threshold can lead to poor sensitivity, especially when there is a large class imbalance (as seen with our training datasets, where there are far more COVID-19 negative cases than positive ones). Thus, we used a grid search to adjust the decision boundary used for identifying COVID-19 positive or negative cases, to improve detection rates at the time of testing. For our purposes, the threshold was optimized to sensitivities of 0.9 to ensure clinically acceptable performance in detecting positive COVID-19 cases. This sensitivity was chosen to exceed the sensitivity of lateral flow device (LFD) tests, which achieved a sensitivity of 56.9% (95% confidence interval 51.7%-62.0%) for OUH admissions between December 23, 2021 and March 6, 2021 [22]. Additionally, the gold standard for diagnosing viral genome targets is by real-time PCR (RT-PCR), which has estimated sensitivities of approximately 80%-90% [29, 30]. Therefore, using a threshold of 0.9 will ensure that models can effectively detect COVID-19 positive cases, and exceed the sensitivities of current diagnostic testing methods.

## 5 Results

### 5.1 Debiasing Ethnicity

After training models on patient cohorts from OUH, we externally validated our models across three external patient cohorts from PUH, UHB, and BH (results shown in Table 4). All models achieved reasonably high AUROC scores across all test sets, comparable to those reported in previous studies (which used similar patient cohorts and features) [6,12,22] (Supplementary Table 10), demonstrating that we trained strong classifiers to begin with. AUROC scores for predicting COVID-19 status stayed relatively consistent across all test sets, achieving the highest performances on the BH cohort (PUH: AUROC range 0.834-0.882 [CI range 0.821-0.893]; UHB: 0.849-0.868 [0.824-0.892]; BH: 0.908-0.923 [0.875-0.954]). With respect to the model used, all models achieved similar AUROCs; however, the highest AUROCs were generally achieved by the XGBoost and NN models (average AUROCs of 0.869, 0.881, 0.885, and 0.881 for RL, adversarial, NN, and XGBoost models, respectively). Using a sensitivity configuration of 0.9, we obtained consistent scores for sensitivity across all models and cohorts (PUH: AUROC range 0.876-0.919 [CI range 0.859-0.933]; UHB: 0.867-0.879 [0.832-0.913]; BH: 0.870-0.935 [0.813-0.976]), with RL achieving the highest sensitivities on the UHB and BH test sets. And, as seen in previous studies, our models achieved high prevalence-dependent NPV scores (*>*0.978), demonstrating the ability to exclude COVID-19 with high-confidence.

**Table 4:** COVID-19 status prediction test results (ethnicity mitigation) across different models and test sets, optimized to sensitivities of 0.9. Results are reported alongside 95% confidence intervals (CIs). Bolded values denoting the best score on each test set is shown for sensitivity and AUROC.

|  |  | Sensitivity | Specificity | PPV | NPV | F1 | AUROC |
| --- | --- | --- | --- | --- | --- | --- | --- |
| PUH | RL | 0.876 (0.859-0.893) | 0.512 (0.506-0.518) | 0.088 (0.083-0.092) | 0.987 (0.985-0.989) | 0.159 | 0.834 (0.821-0.847) |
|  | ADV | 0.879 (0.862-0.896) | 0.595 (0.590-0.601) | 0.104 (0.099-0.109) | 0.989 (0.988-0.991) | 0.186 | 0.865 (0.853-0.877) |
|  | NN | 0.890 (0.874-0.906) | 0.631 (0.625-0.636) | 0.114 (0.108-0.120) | 0.991 (0.989-0.992) | 0.202 | 0.875 (0.863-0.886) |
|  | XGB | **0.919 (0.906-0.933)** | 0.532 (0.526-0.538) | 0.095 (0.090-0.100) | 0.992 (0.991-0.993) | 0.172 | **0.882 (0.871-0.893)** |
| UHB | RL | **0.879 (0.845-0.913)** | 0.574 (0.563-0.584) | 0.079 (0.070-0.087) | 0.991 (0.989-0.994) | 0.144 | 0.849 (0.824-0.875) |
|  | ADV | 0.867 (0.832-0.903) | 0.637 (0.627-0.647) | 0.090 (0.080-0.100) | 0.991 (0.989-0.994) | 0.163 | 0.865 (0.840-0.889) |
|  | NN | 0.873 (0.838-0.908) | 0.667 (0.656-0.677) | 0.098 (0.087-0.108) | 0.992 (0.990-0.994) | 0.176 | **0.868 (0.843-0.892)** |
|  | XGB | 0.867 (0.832-0.903) | 0.585 (0.574-0.595) | 0.080 (0.071-0.088) | 0.991 (0.988-0.993) | 0.146 | 0.854 (0.828-0.879) |
| BH | RL | **0.935 (0.894-0.976)** | 0.691 (0.662-0.720) | 0.296 (0.253-0.339) | 0.987 (0.979-0.995) | 0.449 | **0.923 (0.892-0.954)** |
|  | ADV | 0.877 (0.822-0.932) | 0.779 (0.754-0.805) | 0.356 (0.305-0.407) | 0.979 (0.968-0.989) | 0.506 | 0.912 (0.879-0.945) |
|  | NN | 0.870 (0.813-0.926) | 0.803 (0.778-0.827) | 0.380 (0.326-0.433) | 0.978 (0.968-0.988) | 0.529 | 0.912 (0.879-0.945) |
|  | XGB | 0.928 (0.884-0.971) | 0.644 (0.614-0.673) | 0.266 (0.226-0.305) | 0.985 (0.975-0.994) | 0.413 | 0.908 (0.875-0.942) |

Although predictive performance of the RL model only varied slightly with respect to other models, comparison of the outputs across all test sets was found to be statistically significant (p*<*0.0001, by the Wilcoxon Signed Rank Test).

In terms of fairness, the RL model achieved the best performance overall, achieving either the best or second best equalized odds performances (for both TP and FP SDs) across all external test cohorts, except for the FP SD for PUH (results shown in Table 5). The adversarial model achieved the second best performances overall, usually achieving the best or second best scores for one of TP or FP SD metrics. In general, models with an added debiasing functionality (i.e. RL or adversarial models) demonstrably improved equalized odds, with only a slight trade-off in performance (AUROC decreased between 0.002-0.008 across PUH, UHB, and BH cohorts). Similar results were found when models were optimized to sensitivities of 0.85 (full numerical results in Supplementary Tables 11 and 12), with RL generally achieving the best (or second best) results with respect to equalized odds (followed by adversarial training), demonstrating model consistency across small shifts in the decision threshold.

**Table 5:** Equalized odds evaluation for COVID-19 prediction task (ethnicity mitigation) on external test sets, optimized to sensitivities of 0.9. Results reported as SD of true positive and false positive rates, across all hospital labels. Red and blue values denote best and second best scores, respectively.

|  |  | TP (SD) | FP (SD) |
| --- | --- | --- | --- |
| PUH | RL | **0.047** | 0.037 |
|  | ADV | **0.050** | **0.014** |
|  | NN | 0.066 | **0.028** |
|  | XGB | 0.133 | 0.053 |
| UHB | RL | **0.057** | **0.041** |
|  | ADV | 0.072 | **0.039** |
|  | NN | **0.069** | 0.052 |
|  | XGB | 0.106 | 0.071 |
| BH | RL | **0.030** | **0.010** |
|  | ADV | **<0.001** | 0.045 |
|  | NN | 0.059 | 0.039 |
|  | XGB | 0.107 | **0.036** |

With the same goal of mitigating ethnicity biases, we additionally tested our method on a different classification task - patient discharge prediction. As before, we found that all models achieved reasonably high AUROC scores on the test set, comparable to previously reported benchmarks using the same dataset [31]. AUROC scores ranged from 0.829-0.875 (CI range 0.816-0.886), with the XGBoost model achieving the highest score and RL achieving the lowest (Table 6). However, when optimizing sensitivities to 0.9, RL achieved the best results in terms of sensitivity and equalized odds (Table 7), despite a small trade-off in AUROC. This was also the case when sensitivities were optimized to 0.85 (full numerical results in Supplementary Tables 13 and 14), demonstrating model consistency. The output from the RL model compared to those from other models was found to be statistically significant (p*<*0.0001, by the Wilcoxon Signed Rank Test).

**Table 6:** Patient ICU discharge prediction test results (ethnicity mitigation) across different models and test sets, optimized to sensitivities of 0.9. Results are reported alongside 95% confidence intervals (CIs). Bolded values denoting the best score on the test set is shown for sensitivity and AUROC.

| Model | Sensitivity | Specificity | PPV | NPV | F1 | AUROC |
| --- | --- | --- | --- | --- | --- | --- |
| RL | **0.897 (0.882-0.912)** | 0.539 (0.531-0.547) | 0.171 (0.163-0.179) | 0.980 (0.977-0.983) | 0.287 | 0.829 (0.816-0.841) |
| ADV | 0.885 (0.869-0.901) | 0.637 (0.629-0.645) | 0.205 (0.195-0.214) | 0.981 (0.979-0.984) | 0.333 | 0.861 (0.849-0.873) |
| NN | 0.884 (0.868-0.899) | 0.600 (0.592-0.608) | 0.189 (0.180-0.198) | 0.980 (0.977-0.983) | 0.312 | 0.847 (0.835-0.859) |
| XGB | 0.883 (0.867-0.899) | 0.674 (0.666-0.682) | 0.223 (0.212-0.233) | 0.982 (0.979-0.985) | 0.355 | **0.875 (0.863-0.886)** |

**Table 7:**
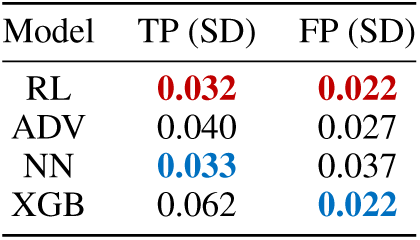
Equalized odds evaluation for patient ICU discharge status prediction task (ethnicity mitigation) on test set, optimized to sensitivities of 0.9. Results reported as SD of true positive and false positive rates, across all hospital labels. Red and blue values denote best and second best scores, respectively.

**Table 8:** COVID-19 status prediction test results (hospital mitigation) across different models, optimized to sensitivities of 0.9. Results are reported alongside 95% confidence intervals (CIs). Bolded values denoting the best score on the test set is shown for sensitivity and AUROC.

|  | Sensitivity | Specificity | PPV | NPV | F1 | AUROC |
| --- | --- | --- | --- | --- | --- | --- |
| RL | **0.887 (0.868-0.906)** | 0.622 (0.614-0.630) | 0.155 (0.146-0.164) | 0.986 (0.984-0.989) | 0.264 | 0.879 (0.865-0.892) |
| ADV | 0.882 (0.862-0.901) | 0.642 (0.634-0.650) | 0.161 (0.152-0.171) | 0.986 (0.983-0.988) | 0.272 | 0.882 (0.869-0.896) |
| NN | 0.879 (0.859-0.898) | 0.676 (0.669-0.684) | 0.175 (0.165-0.185) | 0.986 (0.984-0.989) | 0.292 | 0.891 (0.879-0.904) |
| XGB | 0.875 (0.855-0.895) | 0.720 (0.712-0.727) | 0.196 (0.185-0.207) | 0.987 (0.984-0.989) | 0.320 | **0.900 (0.887-0.912)** |

**Table 9:**
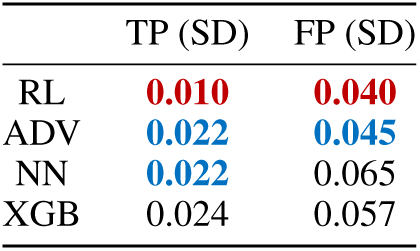
Equalized odds evaluation for COVID-19 status prediction (hospital mitigation) on test set, threshold adjusted to sensitivities of 0.9. Results reported as SD of true positive and false positive rates, across all hospital labels. Red and blue values denote best and second best scores, respectively.

|  | TP (SD) | FP (SD) |
| --- | --- | --- |
| RL | **0.010** | **0.040** |
| ADV | **0.022** | **0.045** |
| NN | **0.022** | 0.065 |
| XGB | 0.024 | 0.057 |

### 5.2 Debiasing Hospital

To demonstrate the need for inter-hospital bias mitigation, we used t-Stochastic Neighbor Embedding (t-SNE) to visualize a low-dimensional representation of all positive COVID-19 cases across the four NHS sites. From Fig. 3, we can see an isolated green cluster corresponding exclusively to a subset of presentations from OUH. This suggests that the training data can be clustered by, and thus, is biased to site-specific features such as annotation methods, data truncation, measuring devices, or collection/processing tools. These distribution shifts emphasize the importance of considering site-specific biases during model development.

**Figure 3:**
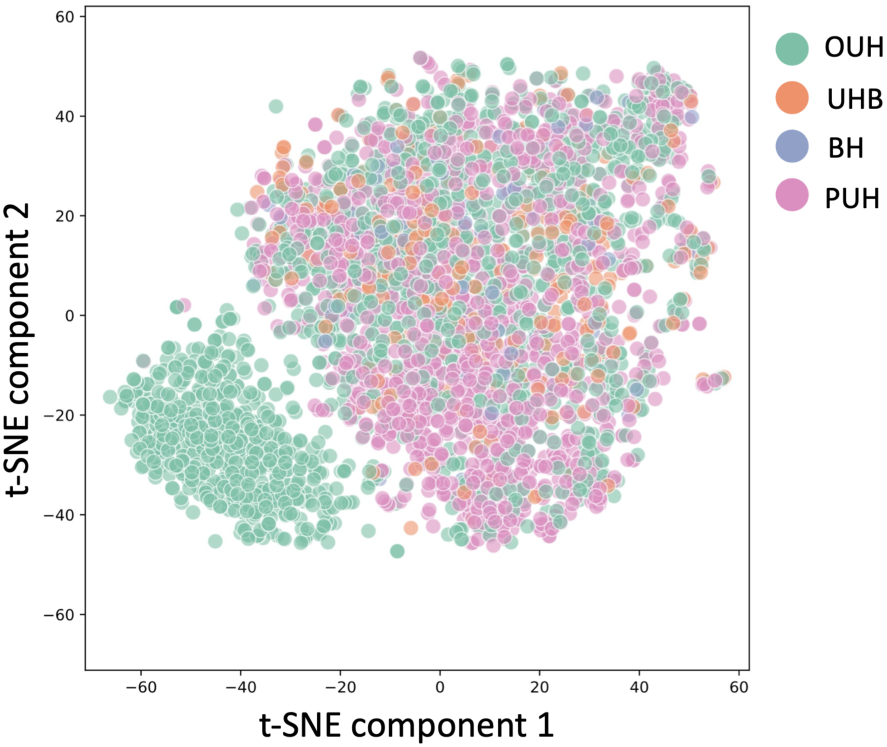
Visualization via t-SNE representation of datasets used in the study, including all positive COVID-19 cases across four NHS trusts (OUH, PUH, UHB, BH).

For this bias mitigation task, we tested all models on a held-out set which included patient presentations from all four hospitals. Model performances were higher than those achieved in the previous COVID-19 status prediction task (focused on ethnicity mitigation); and, as before, although scores were relatively consistent, the XGBoost and NN models achieved the highest AUROC scores (AU-ROCs of 0.879 [CI 0.865-0.892], 0.882 [0.869-0.896], 0.891 [0.879-0.904], and 0.900 [0.887-0.912] for RL, adversarial, NN, and XGBoost models, respectively). Using a sensitivity configuration of 0.9, we obtained consistent scores for sensitivity across all models (sensitivities of 0.887 [0.868-0.906], 0.882 [0.862-0.901], 0.879 [0.859-0.898], and 0.875 [0.855-0.895] for RL, adversarial, NN, and XGBoost models, respectively), with RL achieving the highest sensitivity. And, again, all models achieved high prevalence-dependent NPV scores (*>*0.986), demonstrating the ability to exclude COVID-19 with high-confidence.

Although the overall predictive performance between models was similar, the output from the RL model compared to those from other models was found to be statistically significant (p*<*0.0001, by the Wilcoxon Signed Rank Test).

In terms of bias mitigation, the RL model achieved the most fair performance, achieving the best result with respect to equalized odds (for both TP and FP SD scores). The adversarial model achieved the second best performance. These results were consistent when models were optimized to sensitivities of 0.85 (full numerical results in Supplementary Tables 15 and 16), demonstrating resilience of debiasing to decision thresholds. Thus, as shown in the previous task, models with an added debiasing functionality (i.e. RL or adversarial frameworks) demonstrably improved equalized odds, with only a slight trade-off in predictive performance (greatest AUROC decrease of 0.021 when comparing the RL model with the XGBoost implementation).

## 6 Conclusion and Discussion

As machine learning becomes increasingly prominent for clinical decision-making, it is essential that these technologies do not reflect or exacerbate any unwanted or discriminatory biases. In order for this to be achieved, principles of fairness need to be considered during model development and evaluation. In this study, we introduced a novel method for training fair, unbiased machine learning models, based on a deep reinforcement learning framework. We evaluated it on two complex, real-world tasks - screening for COVID-19 and predicting patient discharge status - while aiming to mitigate site-specific (hospital) and demographic (patient ethnicity) biases. Through comparison of our RL method against current benchmarks and state-of-the-art machine learning models, we found that RL demonstrably improved outcome fairness, while still performing as a strong classifier in general. We appreciate that looking at variations across select hospitals and ethnicities only addresses a fraction of existing inequities in healthcare; however, we hope that the framework and concepts introduced will encourage the use of both deep reinforcement learning and fairness principles, on a diverse range of prediction and debiasing tasks.

For all tasks, we found that the outcomes of the RL models were less biased compared to those with no bias mitigating component. However, although bias generally decreased, our models did not completely fulfill equalized odds requirements. One contributing factor may be that our training datasets, for all tasks, were imbalanced with respect to the sensitive attribute (i.e., a much larger representation of white patients than other ethnicities; much more data was available from OUH and PUH than UHB and BH). As the base network we are using is a neural network, skewed distributions can potentially give inconsistent results. This has previously been observed for adversarial training, as using balanced data was found to have a much stronger effect on fairness outcomes [13]. Thus, if there is sufficient data, future models could benefit from being trained on balanced datasets.

With respect to fulfilling equalized odds requirements, the advantage of using an RL framework was more observable and clear (i.e. noticeable improvements in TP and FP SDs for RL results over other models) for the patient discharge task and the COVID-19 task that involved mitigating inter-hospital biases. This may be due to the larger amount of training data used in those tasks compared to the COVID-19 task with ethnicity debiasing (14,949 patients compared to 43,754 and 49,305 patients for COVID-19 ethnicity, COVID-19 hospital, and ICU patient discharge tasks, respectively). Having a greater amount of training data may have made it easier for models to confidently differentiate between different classes (for both the main task and the sensitive attribute). This was demonstrated by the COVID-19 tasks, as higher predictive performance was achieved for the hospital-site mitigation task than the ethnicity mitigation task, as the hospital-based task utilized a larger training set.

For the main classification tasks (i.e., COVID-19 prediction and ICU patient discharge prediction), we used threshold adjustment to ensure models achieved high sensitivity. This technique is especially effective when there are large label imbalances in the training data, which was true for both tasks we investigated. However, as seen in the t-SNE visualization (Fig. 3), data can be biased by site-specific factors, including data collection/annotation methods, analyzer brands, and population distributions. Because of this, an optimal threshold is biased on the particular dataset that it was derived from; and thus, the threshold used at one site, may not be suitable for new settings with distinct distributions. This may have contributed to the differing sensitivities between test sites used in the COVID-19 task with ethnicity debiasing. Thus, choosing an optimal decision threshold should be further investigated, as it directly affects both classification and fairness metrics (through the shifting of true positive/true negative rates). Furthermore, in clinical settings, it is especially desirable for models to achieve consistent sensitivity (or specificity) scores across different sites. Even if AUROC is consistent, there can still be varying sensitivities/specificities, which can make it difficult for clinicians to rely on the performance characteristic of a model. Thus, future experiments could consider using site-specific thresholds, tailored during a calibration-phase of deployment at each site, in order to standardise predictive performance [12].

Similarly, further consideration should be given to the trade-off between sensitivity and specificity; and additionally, how this can be connected to notions of fairness. For example, for some tasks, high sensitivity may be desirable, as greater harm is caused by false negatives (such as disease diagnosis). Thus, for these tasks true positive parity (equal opportunity) could be used as the fairness metric, as this requires that the probability of a classifier predicting a sample as the positive class is equal across all classes of the sensitive attribute (note, this is the true positive fairness component of the equalized odds measure). Many other fairness metrics also exist (e.g. statistical parity, test fairness); thus, applications should be optimized to definitions of fairness that are most appropriate to each task.

Although we were able to demonstrate the effectiveness of our model on multi-class sensitive features, it is still important to consider whether a demographic-specific/site-specific model versus a multiclass (more generalized) model is best suited for the task. For example, if the purpose of a model is to support patients within a specific hospital care structure, or to predict the risk of a disease that is known to differ significantly between ethnicities, then using personalized models (trained individually on each class) may be the most appropriate choice. However, for tasks requiring very large computational power, training multiple models can be overwhelming for hospitals to implement; thus, using a more generalized model (such as the debiasing framework introduced here) would be beneficial.

Another related limitation is the challenge in understanding the relationships between genetic, social, and behavioral factors in influencing clinical outcomes. As there are increasing amounts and types of data available (imaging, genetic sequencing, etc.), multi-modal analyses and algorithms could be used to understand the underpinnings of human biology and health. By leveraging multiple modes of data, there is more comprehensive information available for training, resulting in a stronger classifier. Even prior to model development, these can be analyzed to gain better understanding of what tasks can be potentially modeled to begin with, and which features are most relevant to creating an accurate and robust model. These analyses will also help determine what biases actually exist and guide how they can best be mitigated.

Finally, bias may exist with regards to missing data points. To address this issue, we used population median imputation, as this was used in related COVID-19 studies involving the same data cohorts. However, the reason for and nature of data missingness may also reflect inherent biases, including differences in access, practice, or recording protocols. Future studies should explore different methods to measure and account for missing data, in case it may convey important information.

## Data Availability

Data from OUH studied here are available from the Infections in Oxfordshire Research Database, subject to an application meeting the ethical and governance requirements of the Database. Data from UHB, PUH and BH are available on reasonable request to the respective trusts, subject to HRA requirements. The eICU Collaborative Research Database is available online. Code and supplementary information for this paper are available online alongside publication.

## Contributions

JY conceived and designed the study. JY wrote the code, performed the analyses, and wrote the manuscript. JY and AS preprocessed the COVID-19 datasets. All authors revised the manuscript.

## Acknowledgements

Thank you to Rasheed El-Bouri for helping with the extraction of the eICU data. Additionally, we express our sincere thanks to all patients and staff across the four participating NHS trusts; Oxford University Hospitals NHS Foundation Trust, University Hospitals Birmingham NHS Trust, Bedfordshire Hospitals NHS Foundations Trust, and Portsmouth Hospitals University NHS Trust. We additionally express our gratitude to Jingyi Wang & Dr Jolene Atia at University Hospitals Birmingham NHS Foundation trust, Phillip Dickson at Bedfordshire Hospitals, and Paul Meredith at Portsmouth Hospitals University NHS Trust for assistance with data extraction.

## Ethics

United Kingdom National Health Service (NHS) approval via the national oversight/regulatory body, the Health Research Authority (HRA), has been granted for development and validation of artificial intelligence models to detect Covid-19 (CURIAL; NHS HRA IRAS ID: 281832).

The eICU Collaborative Research Database (eICU-CRD) is a publicly-available, anonymized database with pre-existing institutional review board (IRB) approval. The database is released under the Health Insurance Portability and Accountability Act (HIPAA) safe harbor provision. The re-identification risk was certified as meeting safe harbor standards by Privacert (Cambridge, MA) (HIPAA Certification no. 1031219-2).

