## Supplementary Material for "Algorithmic Fairness and Bias Mitigation for Clinical Machine Learning: A New Utility for Deep Reinforcement Learning"

#### A Software Packages and Implementation

Models were implemented using Python (v3.6.9). Scikit Learn (v0.24.1) was used for standardization, median imputation, and calculating performance metrics. Performance metrics were calculated using Scikit Learn and manually programmed. XGBoost baseline models were implemented using the XGBoost library (v1.3.3). Neural network baseline models were implemented using Keras (v2.6.0). Reinforcement learning was set up using Tensorflow (v2.6.2). All models were run using an Intel Xeon E-2146G Processor (CPU: 6 cores, 4.50 GHz max frequency).

#### B Model Architectures

**Neural Network Model**: The rectified linear unit (ReLU) activation function was used for the hidden layers and the sigmoid activation function was used in the output layer. For updating model weights, the Adaptive Moment Estimation (Adam) optimizer was used during training.

**XGBoost Model**: XGBoost is a popular ensemble model that has achieved state-of-the-art results on many machine learning challenges. Ensemble methods combine the predictions of multiple models, such that the generalization error is improves (i.e., contribution of individual error from any individual model is lessened). XGBoost in particular, utilizes a boosting technique, where trees are sequentially added and fit to correct for the prediction errors made by previous models. Default settings were used in all experiments.

**Adversarial Debiasing Model**: The adversarial debiasing architecture used consists of two individual networks – a predictor network, $P$, and an adversary network, $A$. $P$ and $A$ are each a multilayer perceptron (MLP). Here, $P$ is trained to predict COVID-19 status (or patient ICU discharge status), given a set of clinical features. Its raw output, $\hat{y}$ – the predicted probability score, and the true label, $y$, are then used as the input to $A$, which tries to predict $z$. For our purposes, $z$ is either hospital site or ethnicity. We use cross-entropy loss (and binary cross-entropy loss when the feature is binary), where $L_P$ represents the loss for $P$ , and $L_A$ represents the loss for $A$. And as introduced in [6], the loss function for $P$ is:

$$L = L_P - \frac{L_P}{L_A} - \alpha L_A \tag{1}$$

For $P$, the sigmoid activation function is used in the output layer; and for $A$, the softmax activation function is used instead.

#### C COVID-19 Data and Preprocessing

**Oxford University Hospitals NHS Foundation Trust (OUH):** We included all patients attending acute and emergency care settings at OUH who received routine blood tests on arrival, considering presentations before December 1, 2019, and thus before the pandemic, as the COVID-19-negative (control) cohort. We considered presentations during the 'first wave' of the UK COVID-19 pandemic (December 1, 2019 to June 30, 2020) with PCR confirmed SARS-CoV-2 infection as the COVID-19-positive (cases) cohort. We excluded patients who opted out of electronic health record (EHR) research and those who did not receive laboratory blood tests or were younger than 18 years of age. Due to incomplete penetrance of testing during the first wave of the pandemic, and imperfect sensitivity of the PCR test, there is uncertainty in the viral status of patients presenting during the pandemic who were untested or tested negative. We therefore selected a pre-pandemic control cohort during training to ensure absence of disease in patients labelled as COVID-19-negative. Clinical features extracted for each presentation included first-performed blood tests, blood gases, vital signs measurements and PCR testing for SARS-CoV-2 (Abbott Architect [Abbott, Maidenhead,

UK], TaqPath [Thermo Fisher Scientific, Massachusetts, USA] and Public Health England-designed RNA-dependent RNA polymerase assays).

**Portsmouth Hospitals University NHS Foundation Trust (PUH):** PUH considered all patients admitted to the Queen Alexandria Hospital, serving a population of 675,000 and offering tertiary referral services to the surrounding region, between March 1, 2020 and February 28, 2021. Confirmatory COVID-19 testing was by laboratory SARS-CoV2 RT-PCR assay, considering any positive PCR result within 48hrs of admission as a true positive.

**University Hospitals Birmingham NHS Foundation Trust (UHB):** UHB considered all patients admitted to The Queen Elizabeth Hospital, Birmingham, between December 01, 2019 and October 29, 2020. The Queen Elizabeth Hospital is a large tertiary referral unit within the UHB group which provides healthcare services for a population of 2.2 million across the West Midlands. Confirmatory COVID-19 testing was performed by laboratory SARS-CoV-2 RT-PCR assay.

**Bedfordshire NHS Foundation Trust (BH):** BH considered all patients admitted to Bedford Hospital between January 1, 2021 and March 31, 2021. BH provides healthcare services for a population of around 620,000 in Bedfordshire. Confirmatory COVID-19 testing was performed on the day of admission by point-of-care PCR based nucleic acid testing [SAMBA-II & Panther Fusion System, Diagnostics in the Real World, UK, and Hologic, USA].

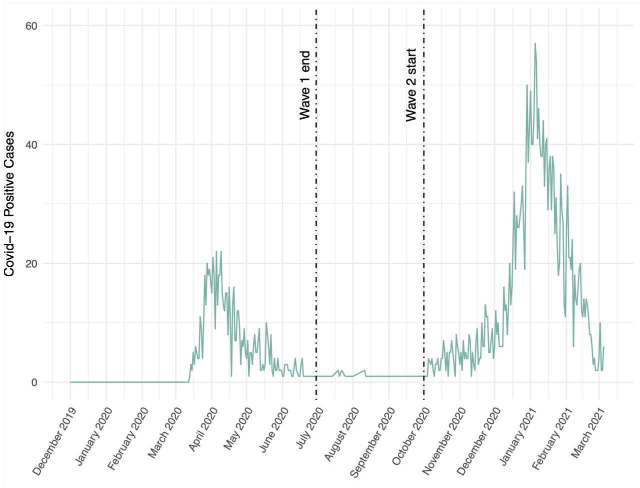


Figure 1: Plot of OUH positive cases, showing the first "wave" of the COVID-19 epidemic in the UK from December 1, 2019 to June 30, 2020; and the second "wave" from October 1, 2020 – March 6, 2021.

Table 1: Summary population characteristics for OUH training cohorts, prospective validation cohort of patients attending OUH, independent validation cohorts of patients admitted to three independent NHS Trusts. *indicates merging for statistical disclosure control.

| | OUH (wave 1 cases) | OUH (wave 2 cases) | PUH | UHB | BH |
|---|---|---|---|---|---|
| ***n*, patients** | 701 | 22,857 | 37,896 | 10,293 | 1177 |
| ***n*, COVID positive** | 701 | 2,012 (8.80%) | 2,005 (5.29%) | 439 (4.27%) | 144 (12.2%) |
| **Sex:** | | | | | |
| - Male (%) | 376 (53.64) | 11409 (49.91) | 20839 (54.99) | 4831 (46.93) | 627 (53.27) |
| - Female (%) | 325 (46.36) | 11448 (50.09) | 17054 (45.0) | 5462 (53.07) | 549 (46.64) |
| Age, yr (IQR) | 72 (55-82) | 67 (49-80) | 69 (48-82) | 63 (42-79) | 68.0 (48-82) |
| **Ethnicity:** | | | | | |
| -White (%) | 480 (68.47) | 17387 (76.07) | 28704 (75.74) | 6848 (66.53) | 1024 (87.0) |
| -Not Stated (%) | 128 (18.26) | 4127 (18.06) | 8389 (22.14) | 1061 (10.31) | $\leq$10 |
| -South Asian (%) | 22 (3.14) | 441 (1.93) | 170 (0.45) | 1357 (13.18) | 71 (6.03) |
| -Chinese (%) | * | 51 (0.22) | 42 (0.11) | 41 (0.4) | $\leq$10 |
| -Black (%) | 25 (3.57) | 279 (1.22) | 187 (0.49) | 484 (4.7) | 36 (3.06) |
| -Other (%) | 34 (4.85)* | 410 (1.79) | 269 (0.71) | 333 (3.24) | 29 (2.46) |
| -Mixed (%) | 12 (1.71) | 162 (0.71) | 135 (0.36) | 169 (1.64) | 13 (1.1) |

#### D eICU-CRD Data and Preprocessing

In terms of clinical applications of AI, patient discharge status has been a popular problem to address, as it can directly influence clinical decision-making, resource allocation, and healthcare costs.

Here, the task was to predict the discharge status of a patient during the course of an ICU stay. Using similar inclusion and exclusion criteria to those used in a previous study [31], we selected adult patients (age > 18) with a minimum of 15 ICU records, and grouped these records into 1 hour windows. We removed any samples that did not have a clear discharge status (i.e., anything that was not "alive" or "expired").

Further preprocessing was performed to remove samples with any missing values, one-hot encode categorical features, and standardize all continuous features to have a mean of 0 and a standard deviation of 1.

We used a 60:20:20 training, validation, and test ratio, resulting in 49,305 training, 16,436 validation, and 16,436 test samples, respectively. As before, the training set was used for model development, hyperparameter selection, and training; the validation set is used for continous validation and threshold adjustment; and after successful development and training, the held-out test set was used to evaluate the performance of the final model.

Table 2: Summary of number of patients, death cases, and hospital case distribution for training, validation, and held-out test set cohorts used in ethnicity debiasing task.

| | Training | Validation | Test |
|---|---|---|---|
| ***n*, patients** | 49305 | 16436 | 16436 |
| ***n*, death** | 4501 | 1486 | 1486 |
| **Ethnicity:** | | | |
| Caucasian (%) | 40285 (81.7) | 13514 (82.2) | 13510 (82.2) |
| African American (%) | 5704 (11.6) | 1832 (11.1) | 1869 (11.4) |
| Hispanic (%) | 2073 (4.2) | 665 (4.0) | 667 (4.1) |
| Asian (%) | 938 (1.9) | 319 (1.9) | 282 (1.7) |
| Native American (%) | 305 (0.6) | 106 (0.6) | 108 (0.7) |

Table 3: Clinical predictors considered for predicting patient discharge status and patient diagnosis.

| Category | Features |
|---|---|
| Demographic features | Gender, age, height, weight |
| Measurements at hospital admission | Non-invasive systolic blood pressure, non-invasive diastolic blood pressure, non-invasive mean arterial pressure, heart rate, supporting oxygen used at admission, blood oxygen saturation, Glasgow coma score, diagnosis at admission |
| Measurements at ICU admission | Glucose |

#### E Hyperparameter Values

Table 4: Hyperparameter values used in COVID-19 status prediction (ethnicity debiasing) task.

| | Hyperparameters: |
|---|---|
| RL | |
| | Gamma = 0.1 |
| | Learning Rate = 0.0001 |
| | Epsilon range = [0.01,1] |
| | Dropout = 0.3 |
| | Hidden nodes = 150 |
| | Optimizer = Adam |
| ADV | |
| | Hidden nodes (predictor) = 100 |
| | Hidden nodes (adversary) = 10 |
| | Dropout = 0.3 |
| | Alpha = 10 |
| | Learning Rate = 0.0001 |
| NN | |
| | Dropout = 0.3 |
| | Hidden nodes = 20 |
| | Learning rate = 0.1 |
| | Optimizer = Adam |
| XGB | |
| | Learning rate = 0.1 |
| | N estimators = 100 |
| | Depth = 3 |

Table 5: Hyperparameter values used in patient ICU discharge status (ethnicity debiasing) task.

| | Hyperparameters: |
|---|---|
| RL | |
| | Gamma = 0.1 |
| | Learning Rate = 0.00009 |
| | Epsilon range = [0.01,1] |
| | Dropout = 0.3 |
| | Hidden nodes = 500 |
| | Optimizer = Adam |
| ADV | |
| | Hidden nodes (predictor) = 200 |
| | Hidden nodes (adversary) = 10 |
| | Dropout = 0.3 |
| | Alpha = 1 |
| | Learning Rate = 0.0001 |
| NN | |
| | Dropout = 0.3 |
| | Hidden nodes = 200 |
| | Learning rate = 0.1 |
| | Optimizer = Adam |
| XGB | |
| | Learning rate = 0.1 |
| | N estimators = 100 |
| | Depth = 3 |

Table 6: Hyperparameter values used in COVID-19 status prediction (hospital-site debiasing) task.

| | Hyperparameters: |
|---|---|
| RL | |
| | Gamma = 0.1 |
| | Learning Rate = 0.00009 |
| | Epsilon range = [0.01,1] |
| | Dropout = 0.3 |
| | Hidden nodes = 100 |
| | Optimizer = Adam |
| ADV | |
| | Hidden nodes (predictor) = 100 |
| | Hidden nodes (adversary) = 10 |
| | Dropout = 0.3 |
| | Alpha = 1 |
| | Learning Rate = 0.0001 |
| NN | |
| | Dropout = 0.3 |
| | Hidden nodes = 10 |
| | Learning rate = 0.1 |
| | Optimizer = Adam |
| XGB | |
| | Learning rate = 0.1 |
| | N estimators = 100 |
| | Depth = 3 |

### F Threshold Values

Table 7: Adjusted Threshold Values Used for COVID-19 status prediction (ethnicity debiasing).

| Model | Threshold | |
|---|---|---|
| | 0.85 | 0.9 |
| RL | 0.50150 | 0.49049 |
| ADV | 0.08417 | 0.05010 |
| NN | 0.07708 | 0.05506 |
| XGB | 0.04204 | 0.01301 |

Table 8: Adjusted Threshold Values Used for patient ICU discharge status prediction (ethnicity debiasing).

| Model | Threshold | |
|---|---|---|
| | 0.85 | 0.9 |
| RL | 0.45445 | 0.44645 |
| ADV | 0.06814 | 0.04609 |
| NN | 0.05105 | 0.02803 |
| XGB | 0.06006 | 0.04304 |

Table 9: Adjusted Threshold Values Used for COVID-19 status prediction (hospital-site debiasing).

| Model | Threshold | |
|---|---|---|
| | 0.85 | 0.9 |
| RL | 0.47347 | 0.46747 |
| ADV | 0.05411 | 0.03206 |
| NN | 0.05405 | 0.03203 |
| XGB | 0.03407 | 0.01804 |

### G Additional Results

#### G.1 Previous Studies

Table 10: Previously published COVID-19 status prediction results. using same datasets and patient cohorts. Sensitivity, specificity, and AUROC shown, alongside 95% confidence intervals, unless otherwise specified.

| Test Set | Sensitivity | Specificity | AUROC |
|---|---|---|---|
| **Soltan et al., 2022.** | | | |
| *Method: XGBoost + SMOTE + Threshold Adjustment (0.9)* | | | |
| OUH | 0.857 (SD 0.009) | 0.686 (SD 0.022) | 0.878 (SD 0.001) |
| PUH | 0.841 (0.825-0.857) | 0.713 (0.709 -0.718) | 0.872 (0.863 -0.882) |
| UHB | 0.788 (0.748-0.824) | 0.747 (0.738 -0.755) | 0.858 (0.838 -0.878) |
| BH | 0.743 (0.666-0.807) | 0.848 (0.825 0. 869) | 0.881 (0.851- 0.912) |
| **Yang et al., 2022.** | | | |
| *Method: Neural Network + SMOTE + ENN + Threshold Adjustment (0.85)* | | | |
| OUH | 0.844 (0.828-0.860) | 0.710 (0.704-0.717) | 0.777 (0.765-0.789) |
| PUH | 0.857 (0.842-0.873) | 0.672 (0.667-0.677) | 0.765 (0.752-0.777) |
| UHB | 0.847 (0.814-0.881) | 0.716 (0.708-0.725) | 0.782 (0.756-0.808) |
| BH | 0.847 (0.789-0.906) | 0.822 (0.799-0.845) | 0.835 (0.793-0.876) |
| **Yang et al., 2022.** | | | |
| *Method: Neural Network + Threshold Adjustment (0.85)* | | | |
| OUH | 0.762 (0.744-0.781) | 0.844 (0.839-0.849) | 0.878 (0.868-0.888) |
| PUH | 0.633 (0.585-0.681) | 0.903 (0.897-0.910) | 0.861 (0.837-0.885) |
| UHB | 0.714 (0.621-0.807) | 0.854 (0.839-0.870) | 0.878 (0.832-0.924) |
| BH | 0.724 (0.561-0.887) | 0.908 (0.869-0.948) | 0.880 (0.798-0.963) |

#### G.2 Debiasing Ethnicity

Table 11: COVID-19 status prediction test results across different models and test sets, optimized to sensitivities of 0.85. Results are reported alongside 95% confidence intervals (CIs).

| | | Sensitivity | Specificity | PPV | NPV | F1 | AUROC |
|---|---|---|---|---|---|---|---|
| PUH | | | | | | | |
| | RL | 0.819 (0.800-0.839) | 0.640 (0.635-0.646) | 0.109 (0.103-0.114) | 0.985 (0.983-0.987) | 0.192 | 0.834 (0.821-0.847) |
| | ADV | 0.829 (0.810-0.849) | 0.698 (0.692-0.703) | 0.128 (0.121-0.135) | 0.987 (0.986-0.989) | 0.222 | 0.865 (0.853-0.877) |
| | NN | 0.848 (0.830-0.867) | 0.704 (0.699-0.710) | 0.133 (0.126-0.140) | 0.989 (0.987-0.990) | 0.230 | 0.875 (0.863-0.886) |
| | XGB | 0.835 (0.816-0.854) | 0.740 (0.735-0.746) | 0.147 (0.139-0.154) | 0.988 (0.987-0.990) | 0.250 | 0.882 (0.871-0.893) |
| UHB | | | | | | | |
| | RL | 0.844 (0.806-0.883) | 0.712 (0.703-0.722) | 0.108 (0.097-0.120) | 0.991 (0.989-0.993) | 0.192 | 0.849 (0.824-0.875) |
| | ADV | 0.824 (0.784-0.864) | 0.757 (0.747-0.766) | 0.123 (0.110-0.136) | 0.990 (0.988-0.993) | 0.214 | 0.865 (0.840-0.889) |
| | NN | 0.830 (0.790-0.869) | 0.741 (0.732-0.750) | 0.117 (0.104-0.130) | 0.991 (0.988-0.993) | 0.205 | 0.868 (0.843-0.892) |
| | XGB | 0.761 (0.716-0.806) | 0.795 (0.786-0.803) | 0.133 (0.118-0.148) | 0.988 (0.985-0.990) | 0.226 | 0.854 (0.828-0.879) |
| BH | | | | | | | |
| | RL | 0.877 (0.822-0.932) | 0.817 (0.793-0.841) | 0.399 (0.344-0.454) | 0.979 (0.970-0.989) | 0.549 | 0.923 (0.892-0.954) |
| | ADV | 0.819 (0.755-0.883) | 0.865 (0.844-0.886) | 0.457 (0.395-0.520) | 0.972 (0.961-0.983) | 0.587 | 0.912 (0.879-0.945) |
| | NN | 0.841 (0.780-0.902) | 0.861 (0.840-0.883) | 0.457 (0.395-0.518) | 0.975 (0.965-0.985) | 0.592 | 0.912 (0.879-0.945) |
| | XGB | 0.848 (0.788-0.908) | 0.845 (0.822-0.867) | 0.432 (0.373-0.491) | 0.976 (0.965-0.986) | 0.572 | 0.908 (0.875-0.942) |

Table 12: Equalized odds evaluation for COVID-19 prediction task on external test sets, optimized to sensitivities of 0.85. Results reported as SD of true positive and false positive rates, across all hospital labels. Red and blue values denote best and second best scores, respectively.

| | | TP (SD) | FP (SD) |
|---|---|---|---|
| PUH | | | |
| | RL | **0.065** | 0.049 |
| | ADV | 0.073 | **0.024** |
| | NN | **0.062** | **0.028** |
| | XGB | 0.207 | **0.028** |
| UHB | | | |
| | RL | **0.066** | **0.039** |
| | ADV | **0.088** | **0.039** |
| | NN | **0.088** | 0.069 |
| | XGB | 0.096 | **0.028** |
| BH | | | |
| | RL | **<0.001** | **0.024** |
| | ADV | **0.051** | 0.030 |
| | NN | 0.053 | 0.035 |
| | XGB | 0.088 | **0.010** |

Table 13: Patient discharge status prediction test results across different models and test sets, optimized to sensitivities of 0.85. Results are reported alongside 95% confidence intervals (CIs).

| Model | Sensitivity | Specificity | PPV | NPV | F1 | AUROC |
|---|---|---|---|---|---|---|
| RL | 0.845 (0.827-0.863) | 0.645 (0.637-0.653) | 0.201 (0.191-0.211) | 0.975 (0.972-0.978) | 0.325 | 0.829 (0.816-0.841) |
| ADV | 0.833 (0.815-0.852) | 0.720 (0.713-0.727) | 0.239 (0.228-0.250) | 0.976 (0.973-0.979) | 0.372 | 0.861 (0.849-0.873) |
| NN | 0.843 (0.825-0.861) | 0.688 (0.681-0.695) | 0.222 (0.212-0.233) | 0.977 (0.974-0.979) | 0.352 | 0.847 (0.835-0.859) |
| XGB | 0.831 (0.813-0.850) | 0.748 (0.741-0.755) | 0.258 (0.246-0.270) | 0.977 (0.974-0.979) | 0.394 | 0.875 (0.863-0.886) |

Table 14: Equalized odds evaluation for patient discharge prediction task on external test set, optimized to sensitivities of 0.85. Results reported as SD of true positive and false positive rates, across all hospital labels. Red and blue values denote best and second best scores, respectively.

| Model | SD TP | SD FP |
|---|---|---|
| RL | **0.042** | **0.017** |
| ADV | **0.056** | **0.020** |
| NN | **0.042** | 0.024 |
| XGB | 0.099 | 0.035 |

##### G.3 Debiasing Hospital

Table 15: Test results across different models, threshold adjusted to sensitivities of 0.85. Results are reported alongside 95% confidence intervals (CIs).

| | Sensitivity | Specificity | PPV | NPV | F1 | AUROC |
|---|---|---|---|---|---|---|
| RL | 0.835 (0.813-0.858) | 0.754 (0.746-0.761) | 0.209 (0.197-0.222) | 0.983 (0.981-0.986) | 0.335 | 0.879 (0.865-0.892) |
| ADV | 0.835 (0.813-0.858) | 0.758 (0.751-0.765) | 0.212 (0.200-0.225) | 0.983 (0.981-0.986) | 0.339 | 0.882 (0.869-0.896) |
| NN | 0.830 (0.807-0.852) | 0.796 (0.789-0.803) | 0.241 (0.227-0.255) | 0.984 (0.981-0.986) | 0.373 | 0.891 (0.879-0.904) |
| XGB | 0.830 (0.808-0.853) | 0.817 (0.811-0.824) | 0.262 (0.247-0.277) | 0.984 (0.982-0.986) | 0.398 | 0.900 (0.887-0.912) |

Table 16: Equalized odds evaluation on test set, optimized to sensitivities of 0.85. Results reported as SD of true positive and false positive rates, across all hospital labels. Red and blue values denote best and second best scores, respectively.

| | TP (SD) | FP (SD) |
|---|---|---|
| RL | **0.014** | **0.033** |
| ADV | 0.022 | **0.037** |
| NN | **0.020** | 0.042 |
| XGB | 0.026 | 0.039 |
